# Fatigue-associated DNA methylation and gene expression profiles differ by disease subtype and activity state in inflammatory bowel disease patients

**DOI:** 10.64898/2026.06.05.26354816

**Authors:** Paula I. Metselaar, Femke Mol, Roni Weiss, Mees J. van der Hoff, Olaf Welting, Wouter J. de Jonge, Peter Henneman, Anje A. te Velde, Mark Löwenberg, Andrew Y.F. Li Yim

## Abstract

**Background and Aims:** Fatigue is a prevalent and disabling symptom in inflammatory bowel disease (IBD), yet its underlying biological mechanisms remain poorly understood. We aimed to characterize fatigue-associated molecular signatures in IBD patients by integrating DNA methylation and mRNA expression analyses.

**Methods:** Peripheral blood was collected from 40 patients with Crohn’s disease (CD), 29 with ulcerative colitis (UC), and 10 healthy controls. Fatigue severity was assessed continuously using the Multidimensional Fatigue Inventory (MFI). Epigenome-wide DNA methylation profiling and mRNA sequencing were performed, identifying differentially methylated regions (DMRs) and differentially expressed genes (DEGs) for active and quiescent CD and UC, adjusting for age, sex, and smoking status. Pathway enrichment analysis was performed on genes with differential methylation and expression.

**Results:** In active CD, more severe fatigue was associated with transcriptional suppression of immune and metabolic pathways (246 DMRs; 1,090 DEGs), versus upregulation of mitochondrial and metabolic processes in quiescent CD (200 DMRs; 1,619 DEGs). In active UC, fatigue was associated with anabolic pathway upregulation and epigenetic silencing of neuroactive pathways (6,927 DMRs; 343 DEGs; 56 concordant genes). Quiescent UC showed transcriptional changes without significant epigenetic pathway enrichment (1,710 DMRs; 3,224 DEGs). Healthy controls exhibited a distinct profile spanning metabolic, immune, and neuronal pathways (8,621 DMRs; 395 DEGs). Fatigue-associated signatures were largely non-overlapping across all five groups.

**Conclusions:** Fatigue-associated molecular profiles differed substantially by disease subtype and activity state, highlighting the biological heterogeneity of IBD-related fatigue and laying the foundation for multi-omics approaches to identify biomarkers and potential therapeutic targets.

## Introduction

Inflammatory bowel diseases (IBD), including Crohn’s disease (CD) and ulcerative colitis (UC), are chronic inflammatory diseases of the gastrointestinal tract that are characterized by flares and periods of quiescent disease. Fatigue represents one of the most commonly reported symptoms among IBD patients in both active and quiescent disease^1–3^. Fatigue has a profound effect on IBD patients by negatively impacting their quality of life, frequently leading to work productivity loss^2–4^. According to a meta-analysis that included 13 studies, pooled prevalence of fatigue was observed in 47% of patients with quiescent and 72% with active IBD^5^. While achieving clinical remission lowers the likelihood of fatigue, a substantial proportion of patients in remission experience persistent fatigue^6,7^. Several hypotheses have been proposed to explain fatigue, including persistent or recurrent active inflammation, residual subclinical inflammation leading to sustained sickness behavior, iron or vitamin deficiencies, unrecognized depression, sleep disturbances, high stress levels and reduced physical activity^8–12^. However, these variables together explain up to 50% of the total variance in fatigue^13^.

Defining fatigue remains a subjective matter. It can be assessed through questionnaires, including the Multidimensional Fatigue Inventory (MFI), Functional Assessment of Chronic Illness Therapy-Fatigue (FACIT-F), visual analogue scales, and the more recently developed IBD fatigue patient self-assessment scale (IBD-F)^14–17^. Few fatigue indices have validated cut-offs for fatigue severity or clinically relevant fatigue^13^. In addition, variability in symptom presentation, overlapping factors such as disease activity, sleep disturbances, depression, and anxiety, and inherent subjectivity of fatigue make accurate measurement challenging^18^. Overall, this underscores the need for molecular markers of fatigue in IBD.

Epigenetics refer to mitotically heritable molecular modifications that affect gene expression without altering the DNA sequence. DNA methylation is a well-characterized epigenetic modification and involves the addition of a methyl group to a base, most often a cytosine within a cytosine-guanine dinucleotide (CpG) context^19^. DNA methylation modulates transcriptional expression depending on the genomic context. Hypermethylation of the gene promoter region is generally associated with transcriptional silencing, while hypomethylation is associated with enhanced gene expression^20^.

Previous epigenetic studies have identified DNA methylation alterations associated with IBD, disease activity, and response to therapies^21–25^, as well as with fatigue in myalgic encephalomyelitis/chronic fatigue syndrome^26–28^ and long COVID^29–33^. Taken together, we hypothesized that DNA methylation may be associated with fatigue in IBD.

To date, no studies have been performed investigating possible associations between DNA methylation, mRNA expression, and IBD-related fatigue. Here, we conducted an exploratory paired multi-omics association study, interrogating the DNA methylome and the transcriptome of peripheral blood obtained from CD and UC patients as well as from healthy controls to identify differences in DNA methylation and gene transcription associated with fatigue severity.

## Methods

### Ethics approval statement

This prospective observational study (NL78138.018.21) was approved by the Medical Ethics Committee of the Amsterdam UMC on December 24, 2021 (METC 2021_207). All participants signed informed consent.

### Study design

IBD patients and healthy controls were recruited at the IBD out-patient clinic of the Amsterdam University Medical Center (UMC) between February 2022 and October 2023. Patients were eligible between 18 and 70 years with a confirmed diagnosis of CD or UC, without pregnancy or serious medical comorbidities. Quiescent CD and UC were defined as a Harvey-Bradshaw Index (HBI) score < 5 or a Simple Clinical Colitis Activity Index (SCCAI) score ≤ 2 for CD and UC, respectively, combined with fecal calprotectin (Fcal) levels < 250µg/ml and/or C-reactive protein (CRP) levels < 5mg/L. Active disease was defined as a HBI score > 5 or SCCAI score > 2, combined with a Fcal level > 250µg/ml and/or CRP level > 5mg/L. At inclusion, patients’ demographic information, medical history, prior and active medication use was documented, clinical activity scores (HBI or SCCAI) were determined, and Fcal, CRP and hemoglobin (Hb) levels were measured in stool and serum samples at the clinical laboratory of the Amsterdam UMC. At this single timepoint, participants also completed three questionnaires and provided blood samples for DNA extraction and mRNA isolation.

### Quantifying fatigue, anxiety, depression and quality of life

Fatigue, anxiety and depression, and quality of life were quantified using the MFI^34^, the Hospital Anxiety and Depression Scale (HADS)^35^, and the 12-item short form health survey (SF-12)^36^, respectively. The MFI score assesses fatigue across 5 dimensions with 20 items rated on a 5-point Likert scale, generating a continuous score between 20 and 100, where higher scores correspond to more severe fatigue. The HADS consists of 7 items per subscale, generating scores between 0 and 21 for both anxiety and depression, where scores ≥8 indicate anxiety and depression. The SF-12 assesses mental and physical quality of life using 12 items. The SF-12 generates T-scores for each component, normed to a healthy reference population (mean = 50), with higher or lower scores indicating better or worse health than the general population.

### Blood collection and preparation

Blood samples were collected in 6.0 ml BD Vacutainer® K2EDTA Tubes for DNA extraction, and 2.5 ml PAXgene® Blood RNA Tubes for mRNA isolation (BD, Franklin Lakes, NJ, USA). Genomic DNA was extracted using QIAsymphony (Qiagen, Valencia, CA, USA) at the Core Facility Genomics, Department of Human Genetics, Amsterdam UMC, according to the manufacturer’s protocol. DNA quantity was assessed using a FLUOstar OMEGA microplate reader. Genomic DNA (750ng) was subjected to bisulfite conversion using the Zymo EZ DNA methylation kit (Zymo Research, Irvine, CA, USA) and subsequent assessment with the Infinium MethylationEPIC v2.0 BeadChip array (Illumina, San Diego, USA) for DNA methylation profiling according to the manufacturer’s protocol^37^. RNA was isolated at the Core Facility Genomics, Department of Human Genetics, Amsterdam UMC. The KAPA mRNA HyperPrep Kit (Roche, Basel, Switzerland) was used for mRNA isolation, cDNA generation, and sequencing adapter ligation. Next generation sequencing was performed on an Illumina NovaSeq X Plus platform with paired-end 150 bp reads at a coverage of 40 million reads per sample.

### Data processing

Analyses were performed within a Snakemake workflow (version 7.25.4) to ensure reproducibility and automated execution. The workflow comprised two independent branches for DNA methylation and mRNA expression analysis. Each branch processed raw data through quality control, normalization, and statistical testing to identify differentially methylated regions (DMRs) and differentially expressed genes (DEGs), respectively. Results were integrated by identifying genes with both differential expression and differential methylation at nominal significance. Visualization and downstream exploratory analyses were conducted in R (version 4.4.0) using *ggplot* (version 3.5.1)^38^ and *ggbio* (version 1.54.0)^39^.

### DNA methylation analyses

Raw data were imported using the Bioconductor package *minfi* (version 1.52.1)^40^ and processed through quality control and functional normalization using *shinyMethyl* (version 1.42.0)^41^. Probes hybridizing to allosomes were excluded from the analysis. M-values were used for statistical analysis and percentage methylation for visualization^42^. Differentially methylated position (DMP) and DMR analyses were performed using *limma* (version 3.62.2)^43^, and *DMRcate* (version 2.12)^44^, respectively. Over representation analysis of DMPs was performed using the gometh() function in the *missMethyl* package (version 1.44.0)^45^.

### mRNA expression analyses

Raw mRNA-sequencing reads were merged per sample and subjected to quality control using *FastQC* (version 0.11.9)^46^ and *MultiQC* (version 1.13)^47^, followed by alignment to reference genome GRCh38 using *STAR* (version 2.7.10a)^48^. Aligned reads were converted to binary files (BAM format) using *SAMtools* (version 1.15.1)^49^. Gene-level counts were obtained with *featureCounts* (version 2.0.1)^50^ using GENCODE v44^51^. Normalization and DEG expression analyses were performed using *DESeq2* (version 1.34.0)^52^, where we regressed against MFI as a continuous variable adjusting for age, sex and smoking. DEGs were analyzed separately for each clinical subset. Gene set enrichment analysis (GSEA) was performed using preranked GSEA from the *fgsea* package (version 1.32.4)^53^. Genes were ranked by their Wald statistic. For genes represented by multiple entries, the mean Wald statistic was used. Gene sets were retrieved from the *msigdbr* package (version 25.1.1, species: Homo sapiens)^54^, using the MSigDB KEGG^55^ subcollection. The minimal gene set size was 15 genes.

### Modelling fatigue

Analyses were performed separately for active and quiescent CD and UC, and healthy (non-IBD) controls. Differential methylation and expression analyses were regressed against MFI scores as a continuous variable adjusting for age, sex, and smoking at time of sampling. Genes and pathways showing nominally significant changes in both methylation and gene expression were visualized. Overlaps across groups were used to identify shared fatigue-associated genes and pathways.

### Clinical statistical analysis

Baseline characteristics were summarized by median and interquartile range (IQR) or counts and percentages. Comparability of patient groups was investigated with Mann-Whitney U tests and Fisher’s exact tests for continuous and categorical variables, respectively. P-values smaller than 0.05 were considered statistically significant. Spearman correlations were calculated between DNA methylation levels (M-values) and corresponding gene expression values.

### Ancillary analyses

Potential confounding by clinical variables was evaluated through Pearson correlations with MFI total score. To assess potential confounding by major depressive disorder, DMRs and associated DEGs identified in this study were compared to DMRs reported in major depressive disorder meta-analyses. DMRs from Li *et al.*^56^ (EPIC array) and Shen *et al.*^57^ (EPIC and 450K) were used, resulting in 14 major depressive disorder-associated DMRs spanning the genes *TNNT3, S100A13, NRXN1, LZTFL1, SS18, GPATCH8, PIKFYVE, CCDC88C, CCDC14, MYO1C, ZNF106, DLEU2, DOT1L,* and *API5*.

Given the incomplete CpG site overlap between EPIC and 450K arrays, overlap was evaluated at the gene-name level.

### Statement on the use of artificial intelligence (AI)

AI assistance (Claude, Anthropic) was used during interpretation of pathway analysis results and manuscript preparation. All content was verified and approved by the authors.

## Results

### Patient characteristics

We included 40 CD patients, 29 UC patients, and 10 healthy controls (**Figure 1A**). Of the IBD patients, 32 had active disease (20 CD, 12 UC) and 37 had quiescent disease (20 CD, 17 UC). All demographic and medical descriptors were balanced in this cohort, except for disease activity indicators and corticosteroid use (**Table 1**). Compared to those with quiescent disease, patients with active disease showed significantly higher HBI (CD) and SCCAI (UC) scores, higher Fcal and CRP levels, higher corticosteroid use, and lower Hb levels. In addition, CD patients with active disease differed significantly from those with quiescent disease in terms of alcohol and recreational drug use, while in the UC group, significant differences were observed in physical quality of life scores. Fatigue, as measured by the MFI, was significantly higher in CD and UC patients compared to healthy controls (both p < 0.001; **Figure 1B**). There was no significant difference in MFI score between active and quiescent disease in either CD (p = 0.343) or UC patients (p = 0.150).

**Figure 1.**
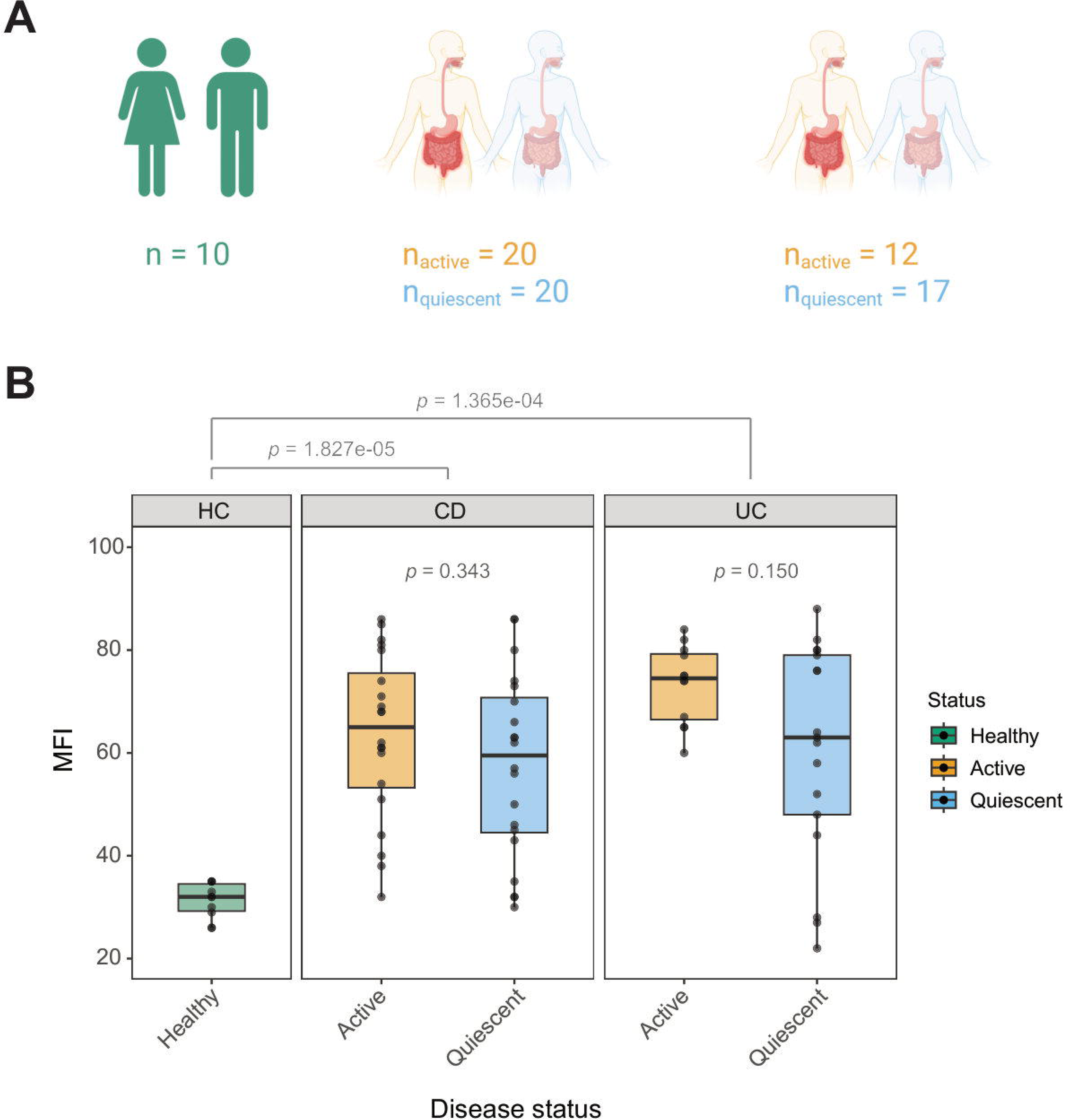
Cohort overview and fatigue distribution across groups. (**A**) Schematic overview of study cohort comprising 3 groups: healthy controls (HC; n = 10), patients with Crohn’s disease (CD; n_active_ = 20, n_quiescent_ = 20), and patients with ulcerative colitis (UC; n_active_ = 12, n_quiescent_ = 17). Disease activity is represented in yellow for active and blue for quiescent disease. (**B**) Boxplots showing the distribution of Multidimensional Fatigue Inventory (MFI) scores across disease types and activity. Boxplots are colored by disease activity (healthy controls: green, active disease: yellow, quiescent disease: blue). Individual datapoints are overlaid on each boxplot; boxes represent interquartile range (IQR) with the median line, and whiskers extend to 1.5 times the IQR; statistical significance (Mann-Whitney U test) is indicated above the relevant comparisons.

**Table 1.** Baseline characteristics of CD patients, UC patients, and healthy controls, grouped by disease activity. P-values were obtained from Mann-Whitney U tests (continuous variables) and Fisher’s exact tests (categorical variables), comparing active vs quiescent disease within each grouping; statistical significance (p < 0.05) is indicated in bold.

| (median [IQR] / n<br>[%]) | Active<br>CD<br>(n=20) | Quiescent<br>CD<br>(n=20) | p-value | Active<br>UC<br>(n=12) | Quiescent<br>UC<br>(n=17) | p-value | HC<br>(n=10) |
| --- | --- | --- | --- | --- | --- | --- | --- |
| CD (n) | 20<br>[100%] | 20<br>[100%] |  |  |  |  |  |
| UC (n) |  |  |  | 12<br>[100%] | 17<br>[100%] |  |  |
| Controls (n) |  |  |  |  |  |  | 10<br>[100%] |
| Sex (female, n) | 13<br>[65%] | 12 [60%] | 1.000 | 8<br>[66.7%] | 7<br>[41.2%] | 0.264 | 5<br>[50%] |
| Age (years) | 48 [28<br>- 54] | 43 [27 -<br>62] | 0.808 | 32 [30 -<br>44] | 36 [27 -<br>51] | 0.690 | 29 [26<br>- 33] |
| BMI (kg/m<sup>2</sup>) | 26 [23<br>- 28] | 27 [23 -<br>30] | 0.537 | 25 [22 -<br>28] | 25 [22 -<br>27] | 0.723 | 22 [21<br>- 24] |
| Active smoking (n) | 3<br>[15%] | 5 [25%] | 0.464 | 2<br>[16.7%] | 3<br>[17.6%] | 1.000 | 0 [0%] |
| Alcohol use (n) | 6<br>[30%] | 13 [65%] | **0.012** | 7<br>[58.3%] | 9<br>[52.9%] | 1.000 |  |
| Recreational drug<br>use (n) | 0 [0%] | 4 [20%] | **0.029** | 1<br>[8.3%] | 3<br>[17.6%] | 0.622 |  |
| Age at diagnosis<br>(years) | 29 [20<br>- 37] | 32 [22 -<br>42] | 0.766 | 26 [18 -<br>34] | 32 [23 -<br>39] | 0.506 |  |
| Disease duration<br>(years) | 10 [8 -<br>16] | 9 [3 - 15] | 0.498 | 6 [3 -<br>14] | 6 [4 - 15] | 0.790 |  |
| Montreal<br>classification (n): |  |  |  |  |  |  |  |
| Montreal A1 | 1 [5%] | 4 [20%] | 0.333 |  |  |  |  |
| Montreal A2 | 15<br>[75%] | 11 [55%] | 0.333 |  |  |  |  |
| Montreal A3 | 4<br>[20%] | 5 [25%] | 0.333 |  |  |  |  |
| Montreal B1 | 11<br>[55%] | 15 [75%] | 0.438 |  |  |  |  |
| Montreal B2 | 5<br>[25%] | 2 [10%] | 0.438 |  |  |  |  |
| Montreal B3 | 4<br>[20%] | 3 [15%] | 0.438 |  |  |  |  |
| Montreal L1 | 3<br>[15%] | 9 [45%] | 0.061 |  |  |  |  |
| Montreal L2 | 4<br>[20%] | 5 [25%] | 0.061 |  |  |  |  |
| Montreal L3 | 13<br>[65%] | 6 [30%] | 0.061 |  |  |  |  |
| Proctitis (E1) |  |  |  | 3 [25%] | 2<br>[11.8%] | 0.158 |  |
| Left-sided UC<br>(E2) |  |  |  | 2<br>[16.7%] | 9<br>[52.9%] | 0.158 |  |
| Pancolitis (E3) |  |  |  | 7<br>[58.3%] | 6<br>[35.3%] | 0.158 |  |
| Current medication<br>(n): |  |  |  |  |  |  |  |
| 5ASA use | 0 [0%] | 1 [5%] | 1.000 | 9 [75%] | 11<br>[64.7%] | 0.694 |  |
| Corticosteroid use | 3<br>[15%] | 0 [0%] | 0.231 | 5<br>[41.7%] | 2<br>[11.8%] | 0.092 |  |
| Immunomodulator<br>use | 2<br>[10%] | 5 [25%] | 0.407 | 2<br>[16.7%] | 3<br>[17.6%] | 1.000 |  |
| Biologic use | 13<br>[65%] | 7 [35%] | 0.113 | 3 [25%] | 5<br>[29.4%] | 1.000 |  |
| Small molecule<br>use |  |  |  | 1<br>[8.3%] | 3<br>[17.6%] | 0.622 |  |
| Antidepressant use<br>(n) | 1 [5%] | 4 [20%] | 0.342 | 2<br>[16.7%] | 3<br>[17.6%] | 1.000 |  |
| Bowel-related<br>surgeries (n): |  |  |  |  |  |  |  |
| Appendectomy | 1 [5%] | 2 [10%] | 1.000 | 1<br>[8.3%] | 2<br>[11.8%] | 1.000 |  |
| Full colectomy | 2<br>[10%] | 1 [5%] | 1.000 |  |  |  |  |
| Ileocecal<br>resection | 6<br>[30%] | 5 [25%] | 1.000 |  |  |  |  |
| Ileostomy | 2<br>[10%] | 2 [10%] | 1.000 |  |  |  |  |
| Partial colectomy | 1 [5%] | 3 [15%] | 0.605 | 1<br>[8.3%] | 0 [0%] | 0.414 |  |
| Proctectomy | 2<br>[10%] | 1 [5%] | 1.000 |  |  |  |  |
| Other surgeries | 0 [0%] | 2 [10%] | 0.487 |  |  |  |  |
| No surgeries | 10<br>[50%] | 7 [35%] | 0.523 | 10<br>[83.3%] | 15<br>[88.2%] | 1.000 |  |
| Duration of disease<br>quiescence<br>(months) | 0 [0 -<br>0] | 11 [0 -<br>29] | **<0.001** | 0 [0 - 0] | 3 [1 - 12] | **<0.001** |  |
| HBI score | 8 [6 - 11] | 2 [0 - 3] | **<0.001** |  |  |  |  |
| SCCAI score |  |  |  | 8 [5 - 9] | 1 [1 - 2] | **<0.001** |  |
| Fcal (µg/g) | 1334 [502 - 4147] | 37 [8 - 130] | **<0.001** | 2351 [963 - 5923] | 60 [29 - 212] | **<0.001** |  |
| CRP (mg/l) | 6 [3 - 17] | 2 [1 - 7] | **0.011** | 7 [1 - 14] | 1 [1 - 2] | **0.020** |  |
| Hb (mmol/l) | 8 [7 - 8] | 9 [8 - 9] | **0.033** | 8 [8 - 8] | 9 [8 - 9] | 0.160 |  |
| SF-12 – physical score | 43 [38 - 51] | 45 [37 - 51] | 0.776 | 41 [33 - 50] | 49 [45 - 53] | **0.032** |  |
| SF-12 – mental score | 40 [32 - 45] | 40 [36 - 50] | 0.441 | 32 [29 - 43] | 38 [34 - 45] | 0.241 |  |
| MFI score | 65 [53 - 76] | 60 [44 - 71] | 0.343 | 74 [66 - 79] | 63 [48 - 79] | 0.150 | 32 [29 - 34] |
| HADS – anxiety score | 7 [4 - 11] | 6 [4 - 11] | 0.532 | 7 [5 - 11] | 3 [2 - 6] | 0.091 |  |
| HADS – depression score | 6 [4 - 9] | 6 [3 - 10] | 0.684 | 8 [5 - 10] | 6 [4 - 11] | 0.706 |  |

5ASA, 5-aminosalicylic acid; BMI, body mass index; CD, Crohn's disease; CRP, C-reactive protein; Fcal, fecal calprotectin;
HADS, hospital anxiety and depression scale; Hb, hemoglobin; HBI, Harvey-Bradshaw index; HC, healthy control; IBD, inflammatory bowel disease; IQR, interquartile range; MFI, multidimensional fatigue inventory; SCCAI, simple clinical colitis activity index; SF-12, 12-item short form health survey; UC, ulcerative colitis.

### Fatigue in active CD associates with suppression of immune recognition pathways, and in quiescent CD with mitochondrial, metabolic, and neurodegeneration pathway activation

In active CD, fatigue modelling yielded 246 significant DMRs, 26,887 DMPs, and 1,090 significant DEGs, of which 4 genes presented differences in both expression and methylation (**Figure 2A**). Transcriptionally, immune recognition pathways (antigen processing and presentation, graft-versus-host disease, autoimmune thyroid disease, allograft rejection), lysosome, and metabolic pathways were suppressed, while ubiquitin-mediated proteolysis, renin secretion, and basal transcription factor pathways were upregulated. On an epigenetic level, no pathways were enriched (**Figure 2C**).

**Figure 2.**
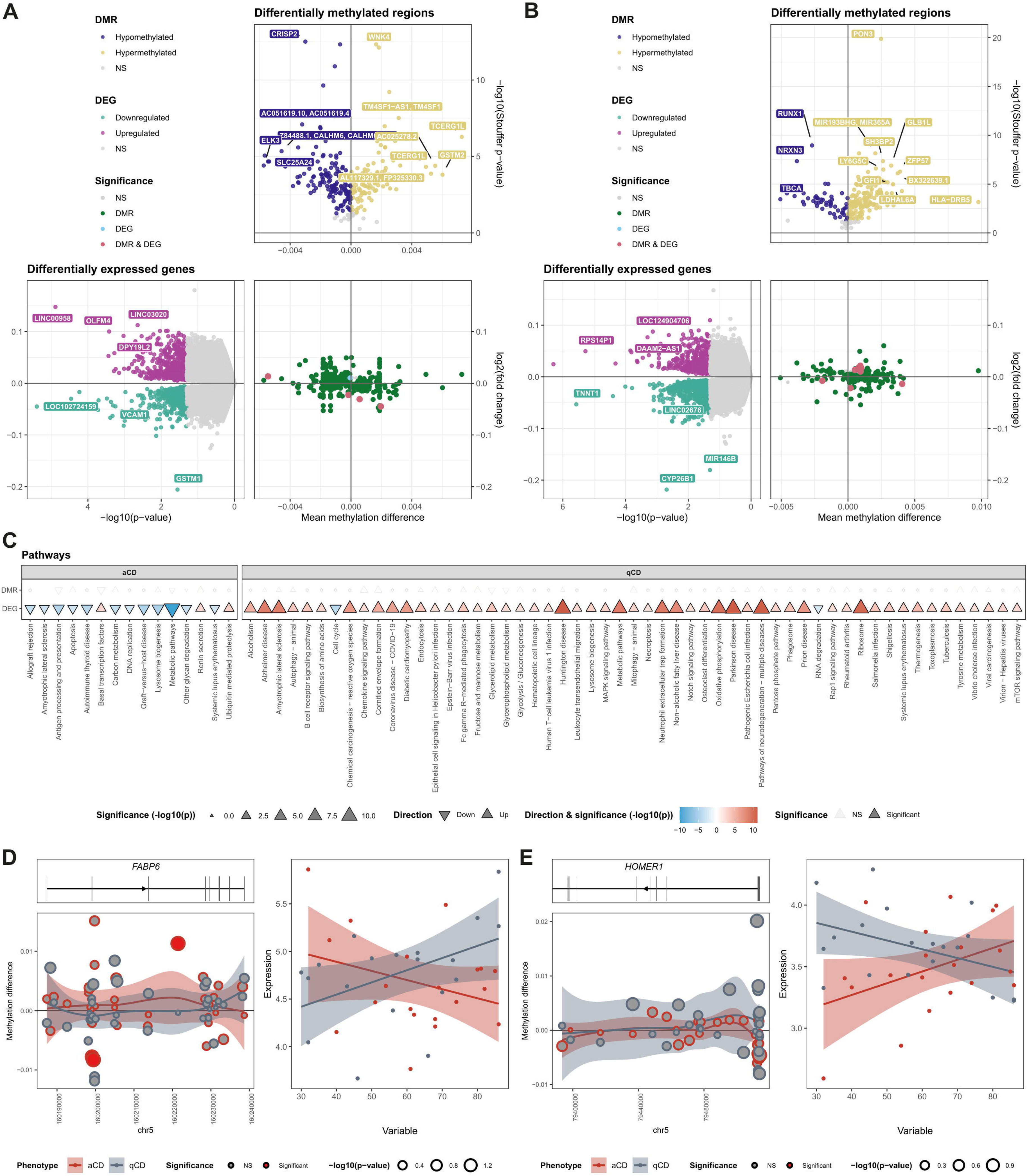
Epigenomic and transcriptomic associations with fatigue in Crohn’s disease. (**A, B**) Volcano plots of differentially methylated regions (DMRs; top-right) and differentially expressed genes (DEGs; bottom-left) in active (**A**) and quiescent (**B**) Crohn’s disease (CD) patients, with an integrated scatter plot (bottom-right) showing the co-occurrence of both modalities. The DMR volcano plot shows mean methylation difference per unit Multidimensional Fatigue Inventory (MFI) on the x-axis and −log_10_(p-value) on the y-axis. The DEG volcano plot shows the log_2_ fold change (LFC) per unit MFI on the y-axis and −log_10_(p-value) on the x-axis. Points are colored by feature type: hypermethylated DMRs (yellow), hypomethylated DMRs (purple), upregulated DEGs (pink), downregulated DEGs (teal), and non-significant (NS) features (grey). Features significant in both modalities (DMR & DEG) are highlighted in red. (**C**) KEGG pathway enrichment plot for DMRs (top row) and DEGs (bottom row) in active and quiescent CD. Pathway direction is indicated by triangle orientation (▴ up, ▾ down). Triangle size and fill intensity both encode −log_10_(p-value), with larger and darker triangles indicating stronger enrichment. Pathways are faceted into 2 columns by disease activity (active CD (aCD) and quiescent CD (qCD)). Within each facet, pathway names are ordered along the x-axis by −log_10_(p-value). (**D, E**) Locus-level integration plots for *FABP6* (**D**) and *HOMER1* (**E**) in active and quiescent CD. For each gene, the left panel shows the genomic track of DMRs across the locus, with individual CpG points sized by −log_10_(p-value) and colored by significance (red: significant, grey: NS). The x-axis represents the coordinates of the locus, and the y-axis shows the methylation difference. The right panel shows a scatter plot of gene expression (normalized counts) against MFI scores for aCD (red) and qCD (grey), with separate linear regression fits and 95% confidence intervals.

Regressing differential methylation and gene expression against MFI scores in quiescent CD yielded 200 significant DMRs, 24,267 DMPs, and 1,619 significant DEGs, where 8 genes presented differences in both expression and methylation (**Figure 2B**). A notable split in pathway activation was observed based on disease activity. While higher fatigue in active CD was associated with transcriptional suppression, higher fatigue in quiescent CD was associated with transcriptional activation. The signal was dominated by upregulation of mitochondrial pathways (oxidative phosphorylation and ribosome), followed by neurodegeneration pathways (Parkinson, Huntington, ALS, Alzheimer, and pathways of neurodegeneration), and global and catabolic activation (pentose phosphate pathway, tyrosine metabolism, biosynthesis of amino acids, glycolysis/gluconeogenesis, and more). Only 2 pathways were downregulated (cell cycle and RNA degradation). As in active CD, no pathways were significantly enriched at an epigenetic level (**Figure 2C**).

Two significant genes with opposing patterns in expression and methylation across disease activity illustrate these signals. *FABP6* (fatty acid-binding protein 6)^58^ was downregulated in active CD (log_2_ fold change (LFC) = −0.026, p = 0.036) and upregulated in quiescent CD (LFC = 0.028, p = 0.021; **Figure 2D**). Conversely, *HOMER1* (Homer scaffold protein 1)^59^, a postsynaptic density scaffolding protein, was upregulated in active CD (LFC = 0.026, p = 0.046) and downregulated in quiescent CD (LFC = −0.026, p = 0.010; **Figure 2E**).

Taken together, fatigue-associated transcriptional changes showed opposite directionality, with no shared pathway directionality between active and quiescent CD.

### Fatigue in active UC associates with anabolic upregulation, immune suppression, and neuroactive epigenetic silencing, and in quiescent UC with a normalized epigenetic landscape

In active UC, regressing differential methylation and gene expression against MFI scores yielded 6,927 significant DMRs, 58,970 DMPs, and 343 significant DEGs, where 56 genes presented significant differences in both expression and methylation (**Figure 3A**). Fatigue severity associated with broad transcriptional upregulation of anabolic pathways (ribosome, ribosome biogenesis, oxidative phosphorylation, DNA replication, RNA degradation, homologous recombination, and N-glycan biosynthesis) and with suppression of immune effector pathways (JAK-STAT signaling, Fcγ R-mediated phagocytosis, osteoclast differentiation, neutrophil extracellular trap formation, and Notch signaling; **Figure 3C**). Within these pathways, *CLEC3B* (tetranectin)^60^, a plasminogen-binding protein involved in extracellular matrix remodeling and immune cell homing, had the largest expression effect size (LFC = −0.187, p = 0.036), supported by a strong promoter-hypermethylated DMR (p = 2.6×10^-8^; Figure 3E).

**Figure 3.**
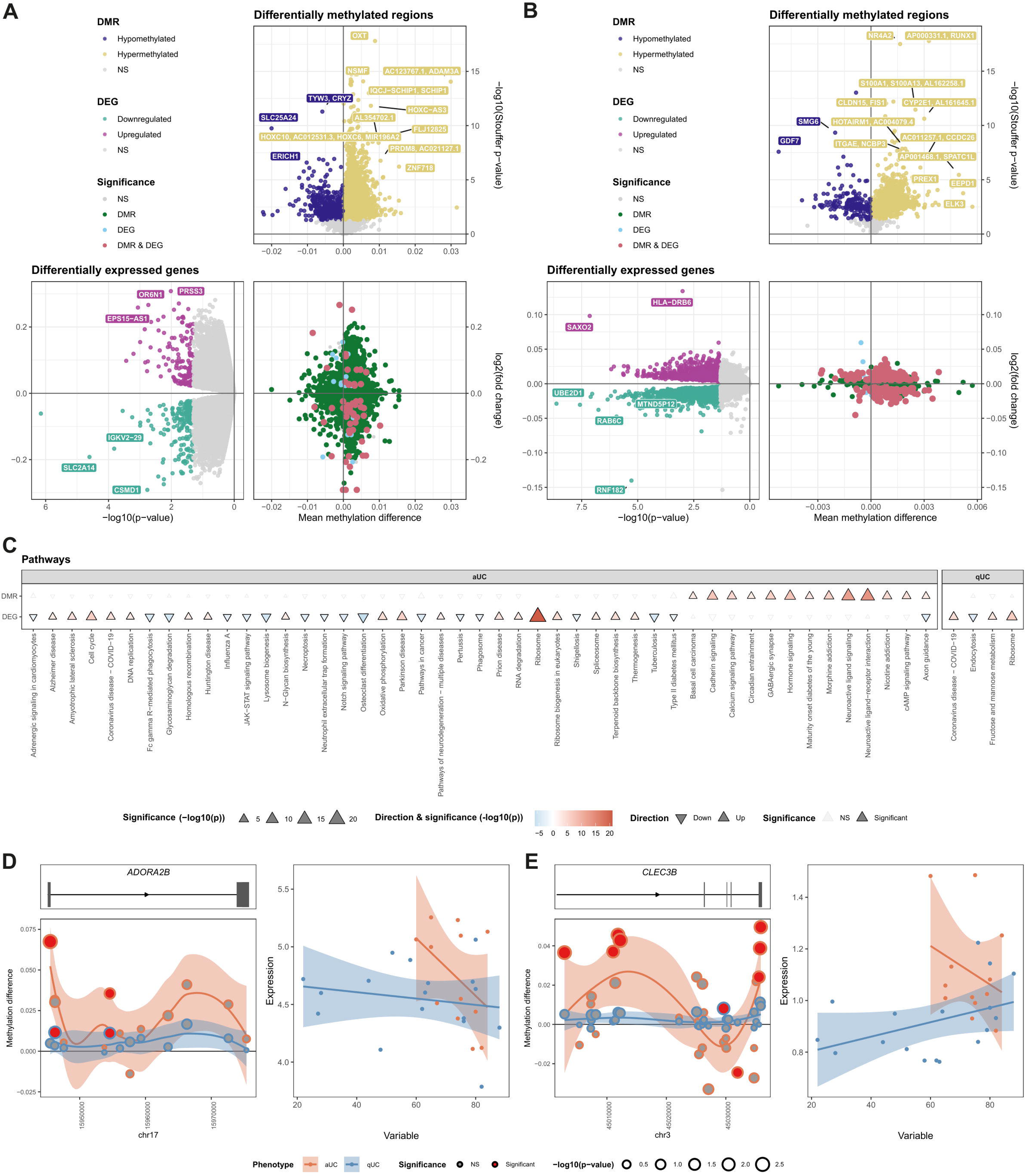
Epigenomic and transcriptomic associations with fatigue in ulcerative colitis. (**A, B**) Volcano plots of differentially methylated regions (DMRs; top-right) and differentially expressed genes (DEGs; bottom-left) in active (**A**) and quiescent (**B**) ulcerative colitis (UC) patients, with an integrated scatter plot (bottom-right) showing the co-occurrence of both modalities. The DMR volcano plot shows mean methylation difference per unit Multidimensional Fatigue Inventory (MFI) on the x-axis and −log_10_(p-value) on the y-axis. The DEG volcano plot shows the log_2_ fold change (LFC) per unit MFI on the y-axis and −log_10_(p-value) on the x-axis. Points are colored by feature type: hypermethylated DMRs (yellow), hypomethylated DMRs (purple), upregulated DEGs (pink), downregulated DEGs (teal), and non-significant (NS) features (grey). Features significant in both modalities (DMR & DEG) are highlighted in red. (**C**) KEGG pathway enrichment plot for DMRs (top row) and DEGs (bottom row) in active and quiescent UC. Pathway direction is indicated by triangle orientation (▴ up, ▾ down). Triangle size and fill intensity both encode −log_10_(p-value), with larger and darker triangles indicating stronger enrichment. Pathways are faceted into 2 columns by disease activity (active UC (aUC) and quiescent UC (qUC)). Within each facet, pathway names are ordered along the x-axis by −log_10_(p-value). (**D, E**) Locus-level integration plots for *ADORA2B* (**D**) and *CLEC3B* (**E**) in active and quiescent UC. For each gene, the left panel shows the genomic track of DMRs across the locus, with individual CpG points sized by −log_10_(p-value) and colored by significance (red: significant, grey: NS). The x-axis represents the coordinates of the locus, and the y-axis shows the methylation difference. The right panel shows a scatter plot of gene expression (normalized counts) against MFI scores for aUC (red) and qUC (blue), with separate linear regression fits and 95% confidence intervals.

At the epigenetic level, increasing MFI scores in active UC were associated with hypermethylation of neuroactive pathways (neuroactive ligand-receptor, neuroactive ligand signaling, GABAergic synapse, calcium signaling, cAMP signaling, hormone signaling, and circadian entrainment). Axon guidance was the only pathway concordantly enriched across both modalities (**Figure 3C**). Illustrative of neuroactive silencing was *ADORA2B* (adenosine A2B receptor)^61^, an anti-inflammatory G protein-coupled receptor, which was hypermethylated (p = 0.009) and transcriptionally downregulated (LFC = −0.088, p = 0.019; **Figure 3D**).

In quiescent UC, 1,710 significant DMRs, 58,225 DMPs, and 3,224 DEGs were identified, of which 271 genes were both differentially expressed and methylated (**Figure 3B**). Transcriptionally, fructose and mannose metabolism, ribosome biogenesis, and coronavirus disease pathways were upregulated, while endocytosis was downregulated (**Figure 3C**). From an epigenetic perspective, no pathways were significantly enriched. However, hypomethylation was observed at loci associated with IBD and with Th1/Th2/Th17 cell differentiation, suggesting partial retention of inflammatory epigenetic memory, though these signals did not remain after multiple testing correction. Both genes illustrative of molecular changes in active UC, *ADORA2B* and *CLEC3B*, showed no signal in quiescent UC (**Figure 3D, E**).

Taken together, active UC was characterized by combined transcriptional and epigenetic signals across anabolic, immune, and neuroactive pathways, whereas quiescent UC showed a largely normalized epigenetic landscape.

### Changes in metabolic, immune, and neuronal pathways reflect baseline molecular variation in healthy controls

To characterize fatigue-associated molecular variation in healthy controls, we performed differential methylation and mRNA expression analyses against MFI scores. This yielded 8,621 significant DMRs, 92,230 DMPs, and 395 significant DEGs (**Figure 4A**). Integrative analysis of these 2 modalities identified 138 DEGs harboring 1 or more MFI-associated DMRs.

**Figure 4.**
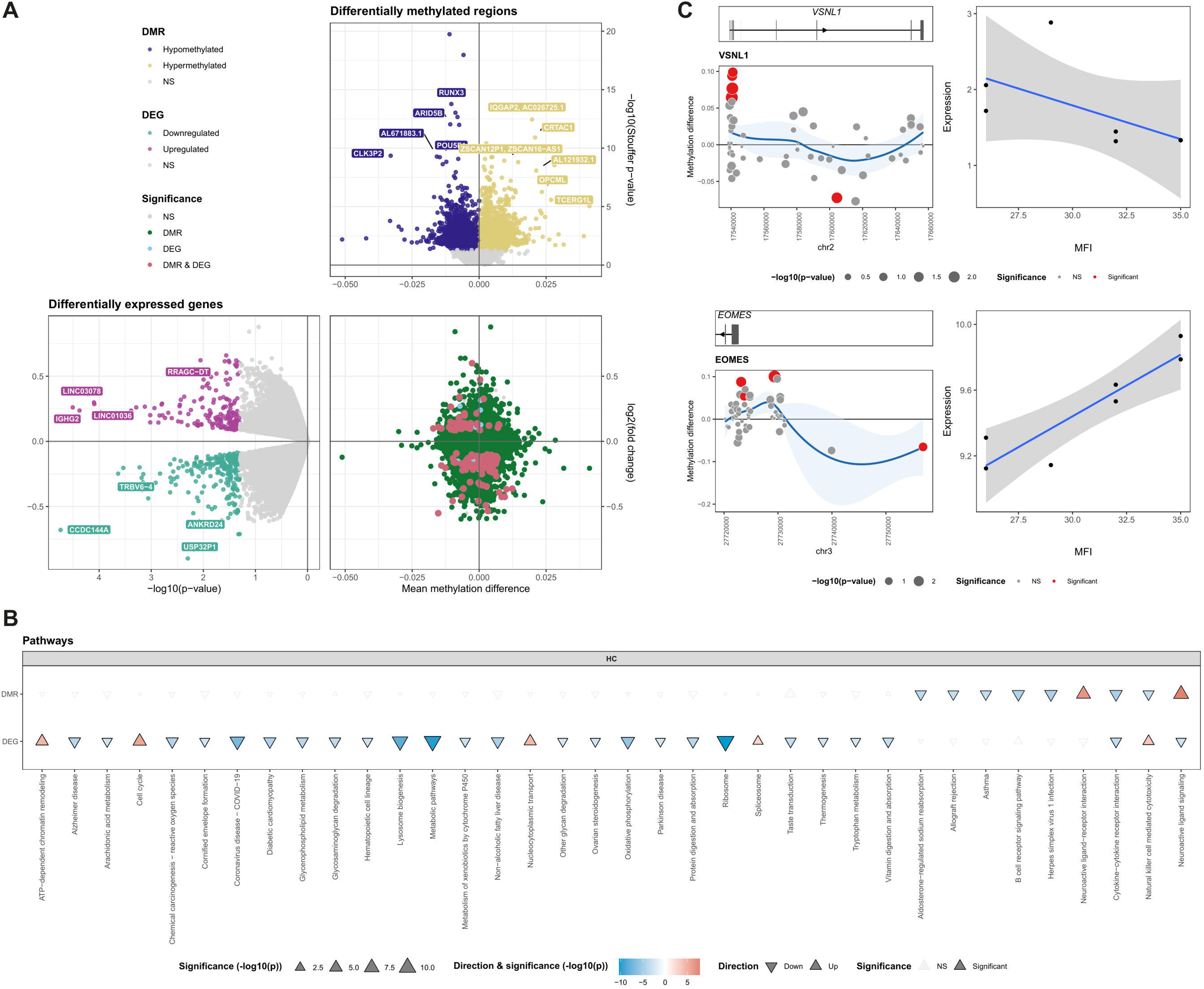
Epigenomic and transcriptomic associations with fatigue in healthy controls. (**A**) Volcano plots of differentially methylated regions (DMRs; top-right) and differentially expressed genes (DEGs; bottom-left) in healthy controls, with an integrated scatter plot (bottom-right) showing the co-occurrence of both modalities. The DMR volcano plot shows mean methylation difference per unit Multidimensional Fatigue Inventory (MFI) on the x-axis and −log_10_(p-value) on the y-axis. The DEG volcano plot shows the log_2_ fold change (LFC) per unit MFI on the y-axis and −log_10_(p-value) on the x-axis. Points are colored by feature type: hypermethylated DMRs (yellow), hypomethylated DMRs (purple), upregulated DEGs (pink), downregulated DEGs (teal), and non-significant (NS) features (grey). Features significant in both modalities (DMR & DEG) are highlighted in red. (**B**) KEGG pathway enrichment plot for DMRs (top row) and DEGs (bottom row) in healthy controls. Pathway direction is indicated by triangle orientation (▴ up, ▾ down). Triangle size and fill intensity both encode −log_10_(p-value), with larger and darker triangles indicating stronger enrichment. Pathway names are ordered along the x-axis by −log_10_(p-value). (**C**) Locus-level integration plots for *VSNL1* (top) and *EOMES* (bottom). For each gene, the left panel shows the genomic track of DMRs across the locus, with individual CpG points sized by −log_10_(p-value) and colored by significance (red: significant, grey: NS). The x-axis represents the coordinates of the locus, and the y-axis shows the methylation difference. The right panel shows a scatter plot of gene expression (normalized counts) against MFI scores for healthy controls, with a linear regression fit and 95% confidence interval.

The transcriptional landscape encompassed a suppressed metabolic compartment (ribosome, lysosome, broad metabolic, oxidative phosphorylation, glycerophospholipid metabolism, protein digestion and absorption, and tryptophan metabolism pathways) alongside an upregulated cluster of nuclear programs (ATP-dependent chromatin remodeling, nucleocytoplasmic transport, cell cycle, and spliceosome) and upregulated natural killer (NK) cell-mediated cytotoxicity (**Figure 4B**).

At an epigenetic level, hypomethylated DMPs were enriched almost exclusively in immune and inflammatory pathways (cytokine-cytokine receptor interaction, B cell receptor signaling, allograft rejection, and asthma). Hypermethylated DMPs were significantly enriched in neuroactive ligand signaling and neuroactive ligand-receptor interaction (**Figure 4B**).

Cross-modal integration revealed hypomethylation and transcriptional upregulation of NK cell-mediated cytotoxicity, exemplified by *EOMES* (eomesodermin)^62^, a master transcription factor governing NK cell terminal differentiation and cytolytic function (DEG LFC = 0.093, p = 0.034 and DMR p = 0.026; **Figure 4C**), and hypermethylation with transcriptional downregulation of neuroactive ligand signaling. Within the latter pathway, *VSNL1* (visinin-like protein 1)^63^, a neuronal calcium sensor protein that modulates receptor recycling, adenylyl cyclase activity, and synaptic plasticity, showed hypermethylation (p = 0.006) and downregulation (LFC = −0.489, p = 0.039; **Figure 4C**). Cytokine-cytokine receptor interaction presented both promoter hypomethylation and transcriptional downregulation (**Figure 4B**).

Taken together, in healthy controls, fatigue-associated methylation and expression changes spanned metabolic, immune, and neuronal pathways, with fatigue scores remaining within a narrow and low range.

### Fatigue-associated molecular profiles are distinct between CD, UC, and healthy controls

Comparing fatigue-associated profiles across the various modalities, each of the 5 groups (active CD, quiescent CD, active UC, quiescent UC, healthy controls) presented a distinct profile, both at the level of individual DMRs and DEGs and at the pathway level (**Figure 5A-C**), indicating that the molecular correlates of fatigue do not follow a single conserved signature. Instead, group-specific patterns emerged.

**Figure 5.**
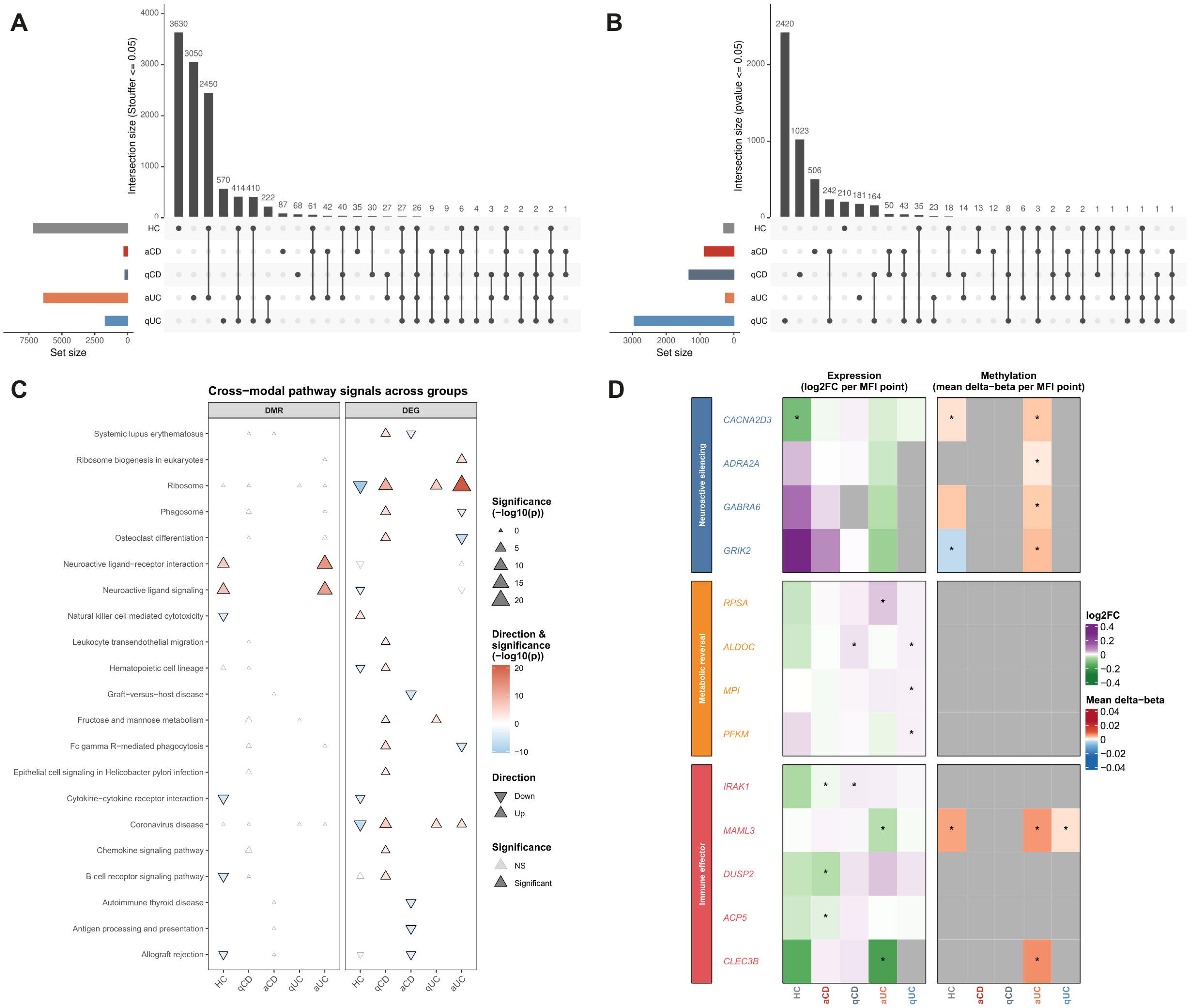
Cross-group integration of epigenetic and transcriptomic associations with fatigue. (**A**) UpSet plot depicting intersections of significant differentially methylated regions (DMRs; Stouffer p < 0.05) across all 5 groups (healthy controls (HC), active ulcerative colitis (aUC), quiescent UC (qUC), active Crohn’s disease (aCD), quiescent CD (qCD)). Set size (left bar chart) indicates total number of significant DMRs per group. Intersection sizes (top bar chart) show the number of DMRs shared across the indicated combination of groups. (**B**) UpSet plot depicting intersections of significant differentially expressed genes (DEGs; p < 0.05) across all 5 groups, structured as described for panel A. (**C**) Cross-modal KEGG pathway enrichment plot for DMRs and DEGs. Pathway direction is indicated by triangle orientation (▴ up, ▾ down). Triangle size and fill intensity both encode −log_10_(p-value), with larger and darker triangles indicating stronger enrichment. Pathways are faceted into 2 columns by modality (DMR and DEG). Within each facet, disease groups are ordered along the x-axis (HC, qCD, aCD, qUC, aUC) and pathways along the y-axis. (**D**) Heatmap of per-gene Multidimensional Fatigue Inventory (MFI) associations with significant DMR and/or DEG signals across groups for 13 selected genes, grouped into 3 functional clusters: neuroactive silencing, metabolic reversal, and immune effector. Left panels show expression of log_2_ fold change (log2FC) per unit MFI; right panels show mean methylation difference (Δβ) per unit MFI. Asterisks (*) indicate statistically significant associations (p < 0.05). Color scales are centered at zero.

At the pathway level, ribosome and coronavirus disease pathways were transcriptionally upregulated in active UC, quiescent UC, and quiescent CD, and downregulated in healthy controls (exemplar gene *RPSA*; **Figure 5D**). This was the only signal shared across all 4 IBD groups, with directionality opposite to healthy controls. Active CD and active UC both showed suppression of immune compartments with increasing fatigue (*IRAK1*, *MAML3*; **Figure 5D**), but engaged different immune branches: active CD showed reduced T cell activation and dendritic cell function (*DUSP2*, *ACP5*), whereas active UC showed reduced monocyte, macrophage, and NK effector activity (*CLEC3B*; **Figure 5D**). Quiescent CD and UC uniquely shared upregulation of fructose and mannose metabolism, implicating mucosal repair biology (*ALDOC*, *MPI*, *PFKM*; **Figure 5D**). Neuroactive pathways were a hallmark of active UC, exemplified by hypermethylation of neuroactive receptor genes (*CACNA2D3*, *ADRA2A*, *GABRA6*, *GRIK2*; **Figure 5D**), and were absent from all other groups. Healthy controls displayed the most distinct profile overall, with more than 17 pathways enriched at the epigenetic or transcriptional level that were absent from all disease groups.

### Fatigue-associated differentially methylated and expressed genes showed no overlap with depression-associated loci

Associations between clinical variables and fatigue severity were explored. Among all factors tested, only depression (HADS) showed a statistically significant positive correlation with fatigue (MFI), with a Pearson correlation coefficient of 0.79 (R^2^ = 0.63, p = 5.5×10^-16^), indicating that higher levels of depression were associated with more severe fatigue (**Supplemental Figure S1**). Subsequently, overlap was assessed between genes associated with major depressive disorder and fatigue in IBD by comparing DEGs harboring DMRs from the current study with 14 previously found DMRs associated with major depressive disorder. No overlap was found between genes that were both differentially expressed and methylated in our dataset and previously reported major depressive disorder-associated DMRs.

## Discussion

In this exploratory multi-omics study, we identified genes exhibiting concomitant differential DNA methylation and gene expression associated with fatigue severity in CD and UC. By integrating DNA methylation and transcriptomic data, we provide a molecular characterization of fatigue as a biologically heterogeneous phenotype that differs not only between disease type (CD vs UC), but also by disease activity (active vs quiescent disease). To our knowledge, this is the first study to jointly assess DNA methylation and mRNA expression in relation to fatigue in both CD and UC.

In CD, fatigue corresponds to two distinct molecular states, depending on disease activity. In active CD, more severe fatigue was associated with transcriptional suppression of immune recognition pathways and metabolic processes, consistent with a state of immune exhaustion during ongoing inflammation^64^. In contrast, in quiescent CD, fatigue was characterized by transcriptional upregulation of mitochondrial, metabolic, and neurodegeneration-related pathways, suggesting a persistent, metabolically active state despite quiescence. This pattern parallels the persistent activation of chronic inflammatory pathways described in post-acute infection syndromes (i.e., long COVID)^65^. Notably, these opposing transcriptional profiles were not mirrored at the epigenetic level, where adaptive immune loci remained accessible across both disease states. This dissociation suggests that DNA methylation reflects a more stable “epigenetic memory” of prior immune activation, whereas gene expression captures the current functional state^66,67^.

These findings imply that fatigue in CD is not a single biological entity but rather encompasses at least two mechanistically distinct states that converge into a similar patient-reported phenotype. In active CD, fatigue may arise from immune and metabolic exhaustion in the context of sustained inflammation, a concept aligned with the broader framework of sickness behavior driven by pro-inflammatory cytokines^68^. In contrast, in quiescent CD, fatigue may reflect incomplete resolution of inflammation, with persistent metabolic activation and immune sensitivity.

In UC, fatigue was associated with distinct molecular mechanisms that differed from CD. In active UC, fatigue severity was linked to transcriptional upregulation of anabolic pathways, alongside suppression of immune effector functions. At the epigenetic level, this state was characterized by widespread hypermethylation of neuroactive ligand-receptor and signaling pathways. The detection of neuroactive signals in peripheral blood was initially unexpected, given that DNA methylation was measured in genomic DNA from whole blood. However, blood transcriptomes recover a substantial fraction of brain-expressed transcripts, and peripheral immune cells intrinsically express a broad range of neurotransmitter receptors that modulate their function and trafficking^69,70^. Methylation changes at these loci may therefore reflect altered neuroimmune signaling within the immune compartment itself.

Given that neurotransmitter receptors are increasingly recognized as modulators of immune cell function and systemic inflammation^71^, epigenetic silencing of these pathways in immune cells may reflect altered neuroimmune communication contributing to fatigue. In contrast, quiescent UC exhibited a largely normalized epigenetic landscape, with residual but non-significant signals suggesting partial retention of inflammatory memory.

In healthy controls, fatigue scores were consistently and significantly lower compared to CD and UC patients, with a narrow distribution. Consequently, molecular associations identified in this group could reflect baseline variation rather than fatigue-specific biology. This is supported by the prominence of metabolic, immune, and neuronal signaling pathways, which are well-recognized components of physiological variability^72,73^. The distinct profile observed in healthy controls compared to CD and UC patients further reinforces that fatigue in IBD patients reflects disease-associated processes rather than physiological variation.

Between the different groups, overlap in fatigue-associated molecular signatures was remarkably limited. Only a small number of DMRs and DEGs were shared between disease states, indicating that fatigue represents a context-dependent phenotype, shaped by disease subtype and inflammatory activity. The ribosome and coronavirus disease pathways were among the few shared signals, displaying opposite directionality between CD and UC patients and healthy controls. Alterations in these pathways have been linked to cellular energy metabolism and fatigue in myalgic encephalomyelitis/chronic fatigue syndrome^74,75^.

Several additional observations merit consideration. First, the magnitude of molecular signal varied substantially between the different groups, with particularly strong signals in active UC and healthy controls. While this may reflect biological differences, it may also be influenced by variability in fatigue distribution and sample size between the different groups. Second, the greater number of transcriptionally enriched pathways in quiescent CD compared to active CD, despite similar numbers of overlapping methylation and expression signals, suggests a broader, but potentially less coordinated transcriptional response in remission. The opposite pattern was observed in UC, where broader transcriptional and epigenetic signals were observed in active disease, while quiescent UC presented a more limited epigenetic landscape.

Depression was strongly associated with fatigue severity, consistent with prior literature demonstrating substantial overlap in symptomatology and clinical burden^18^. However, no overlap was observed between fatigue-associated genes (both differentially methylated and expressed) in the present study and previously reported methylation signatures of major depressive disorder derived from non-IBD psychiatric cohorts. This lack of overlap suggests that the identified signals are specific to fatigue in IBD and do not merely reflect underlying depressive symptoms, indicating that fatigue and depression are underpinned by distinct molecular mechanisms despite their clinical co-occurrence^11^.

Our study has several strengths and limitations. A key methodological strength is the integration of two molecular layers, focusing specifically on genes showing concordant signals in methylation and expression. While both DMRs and DEGs were identified at nominal significance, their overlap provides an additional level of biological evidence, linking epigenetic variation to functional transcriptional differences. All analyses were adjusted for established determinants of DNA methylation, including age, sex, and smoking status^76–78^. Stratification by CD and UC activity further enabled disentangling fatigue-associated mechanisms in active versus quiescent disease. Limitations include the relatively small number of patients with low fatigue scores, particularly in UC, which limited our ability to capture the full fatigue spectrum. The use of bulk whole blood obscures cell type-specific effects, as DNA methylation and gene expression are cell type-dependent^79^. The reliance on nominal significance thresholds increased sensitivity for integrative analyses but may have introduced false positive results, emphasizing the need for validation. Finally, fatigue measured by the MFI carries inherent subjectivity, and inter-individual differences in how patients perceive and report fatigue may have contributed to variability in molecular signal magnitude across the different groups.

Future studies should validate these findings in larger and more heterogeneous cohorts, particularly with greater representation of non-fatigued patients. Longitudinal study designs would help establish whether the observed epigenetic and transcriptional changes precede or follow the development of fatigue. Furthermore, advances in the objective measurement of fatigue beyond self-report questionnaires remain an important unmet need. The development of validated biological markers of fatigue severity would strengthen future multi-omics investigations. Finally, single-cell approaches may further resolve cell type-specific mechanisms underlying fatigue in IBD.

In conclusion, the present study demonstrates that fatigue in CD and UC patients is associated with distinct and largely non-overlapping molecular signatures across disease subtypes and activity states. By integrating DNA methylation and gene expression analyses, we show that fatigue is not a uniform biological entity, but likely reflects context-dependent immune, metabolic, and neuroimmune processes. These findings provide a framework for future mechanistic studies and may lay the foundation for developing diagnostic and therapeutic strategies to address fatigue in IBD.

## Supporting information

Supplemental Figure S1. Correlation between fatigue and depression in inflammatory bowel disease patients

## Data Availability

Summary statistics from the DNA methylation and RNA-seq analyses are publicly available on Zenodo. Analysis code and pipelines are available on Github.

https://zenodo.org/uploads/20325074

https://github.com/fmol98/multiomic-IBD-fatigue/tree/main

## Acknowledgements

We thank all participants for their contribution to this study.

## Data Availability Statement

Summary statistics from the DNA methylation and RNA-sequencing analyses are publicly available on Zenodo at <u>10.5281/zenodo.20325074</u>. Analysis code and pipelines are available on Github at <u>fmol98/multiomic-IBD-fatigue</u>.

## Authors’ contributions

PIM, AAtV, ML, AYFLY contributed to the conception and design of the study; PIM, MJvdH, OW, ML contributed to acquisition of data; PIM, FM, RW, PH, ML, AYFLY contributed to analysis and interpretation of data. All authors contributed to drafting the article and revised it critically for important intellectual content. All authors gave their final approval of the manuscript.

