## Supplemental Figure S1. Correlation between fatigue and depression in inflammatory bowel disease patients for "Fatigue-associated DNA methylation and gene expression profiles differ by disease subtype and activity state in inflammatory bowel disease patients"

**Supplementary material**

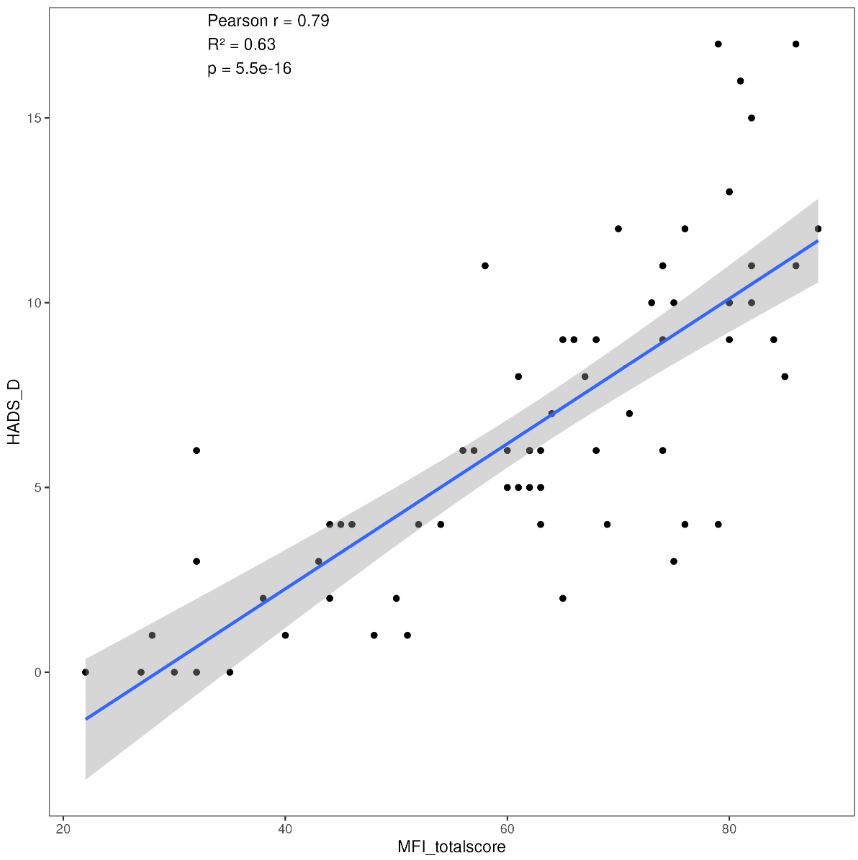

**Supplemental Figure S1. Correlation between fatigue and depression in inflammatory bowel disease patients**

Scatter plot showing the Pearson correlation between Hospital Anxiety and Depression Scale – Depression subscale (HADS-D) scores (y-axis) and Multidimensional Fatigue Inventory (MFI) scores (x-axis) across all inflammatory bowel disease patients, with a linear regression line and shaded 95% confidence interval.
